# Development of the IcanSDM scale to assess primary care clinicians’ ability to adopt shared decision-making

**DOI:** 10.1101/2020.07.01.20144204

**Authors:** Anik MC Giguere, Laura-Mihaela Bogza, Laetitia Coudert, Pierre-Hugues Carmichael, Jean-Sébastien Renaud, France Légaré, Anja Lindig, Philippe Voyer

## Abstract

**Introduction:** Implementation of shared decision making (SDM) remains a challenge. To support implementation studies, we sought to develop and validate the IcanSDM scale that assesses clinicians’ perceptions of their ability to adopt SDM.

**Methods:** An expert panel reviewed the literature on clinician-reported barriers to SDM adoption, to create an 11-item preliminary scale. A convenience sample of 16 clinicians from Québec (Canada) completed the IcanSDM and the Belief about Capabilities subscale of the CPD-Reaction instrument (BCap), before and after SDM training. We audio-recorded their comments as they completed the scale. We measured IcanSDM’s internal consistency, sensitivity to change and correlation with BCap. Partial correlation coefficients and item analyses suggested removing three items. We then tested the 8-item IcanSDM with a new sample of 17 clinicians.

**Results:** In the 11-item IcanSDM version, three items lacked clarity or responsiveness, or showed negative partial correlations with the whole instrument. We thus removed these items. The revised 8-item version gave Cronbach’s alphas of 0.63 before and 0.71 after training, and a 16% improvement in IcanSDM total score after training, compared to before training (p<0.0001). We also found a significant correlation between IcanSDM and the BCap before training (p=0.02), but not after (p=0.46).

**Discussion:** IcanSDM is the only instrument measuring this construct. It could thus help bridge the gap in our ability to understand the determinants of clinicians’ SDM behavior intentions and thus help improve SDM implementation impacts and efforts. IcanSDM requires testing with a larger sample to confirm its responsiveness.

**Lessons for practice:**

- IcanSDM assesses clinicians’ perceived ability to adopt shared decision making.
- IcanSDM demonstrated adequate validity and reliability but needs more testing to confirm its responsiveness.
- IcanSDM is promising to assess the impacts of training in shared decision making and other initiatives to implement shared decision making.

## Introduction

The pinnacle of patient-centered care, shared decision-making (SDM) is defined as a process whereby patients and clinicians collaborate to make choices about patient health.^1^ During this journey, the clinician provides information on the options and research-based outcomes relevant to their health status, to ultimately help them clarify and incorporate their preferences and values into decision making.^2–6^ This approach can be facilitated by patient decision aids that have been shown to help improve patient knowledge, accuracy of risk perception, congruency between informed values and care choices, and satisfaction.^7^ Nevertheless, a review of 33 studies demonstrated that SDM is regularly overlooked during clinical encounters, or only partially used.^8^

While an ever growing number of countries are experimenting with and committing to delivering patient-centered care through SDM, implementation in daily practice remains a challenge.^9–11^ Despite the fact that clinicians mostly agree with the principles of SDM,^12^ they still do not use it in their daily practice.^8^ Current implementation efforts frequently rely on the distribution of patient decision aids by clinicians, and on interventions targeting clinicians, such as educational meetings.^13^ However, clinicians still report a number of barriers to distributing decision aids, and intensive implementation efforts are only partially successful.^14^

With the wealth of research studying barriers to implementation, interventions are now tailored to limit those barriers. For example, to guide clinicians around and beyond the perceived barrier of SDM being difficult to put into use, several training programs in SDM are being implemented.^15^ Clinicians can follow these types of training programs during the pre- and post-licensure phases to gain general knowledge on SDM, understand what it involves, and learn how to use it properly during their clinical encounters.^15,16^ Depending on the training program, some research findings report increased SDM after training.^13^ However, the barriers are only assessed qualitatively, and few measures are available to study the individual factors underlying clinicians’ adoption of SDM.

A recent systematic review inventoried and appraised 40 scales to measure SDM processes, which are the observed and perceived processes during the deliberation phase.^17^ An earlier and broader review described only a few scales to measure the two other SDM domains, which are decision antecedents and decision outcomes.^18^ Moreover, the scales to assess decision antecedents assess patients’−and not clinicians’− preparedness for decision making, such as autonomy preference, decision self-efficacy, and patient attitudes and beliefs.^18^ To the best of our knowledge, there are no scales for evaluating clinicians’ perceptions of the barriers to SDM, despite the fact that these perceptions are key determinants of SDM implementation.^19^

Among the diverse types of barriers to adopting SDM, clinicians consistently report time constraints^19,20^ or the lack of applicability to specific clinical situations or patients.^20^ The Theory of Planned Behaviour^21^ suggests that beliefs about resources and opportunities are indicative of perceived behavioural control and one of three key elements that influence behaviour intention and, ultimately, actual behaviour. A systematic and quantitative assessment of the extent to which clinicians perceive these barriers would therefore be helpful to identify potential training needs and assess training and the effectiveness of program implementation, but such a tool does not yet appear to be available for SDM.^22^

We therefore sought to develop and validate a new scale, the IcanSDM scale to assess clinicians’ perceptions of their ability to adopt SDM. IcanSDM is a self-report scale completed by clinicians. It may be used to evaluate the impacts of training or program implementation, or to help tailor these programs to maximize their impacts. While the current validation study included clinicians who work in primary care settings, IcanSDM is intended for clinicians of any profession working in any setting, as healthcare is increasingly provided by interprofessional teams.^15^

This paper presents the first steps taken to develop and validate this scale.

## Methods

### Study Design

This is a secondary study conducted during a larger study to develop e-TUDE, a professional distance-training program on SDM. Details on the user-centered and theory-based development and evaluation of e-TUDE are reported elsewhere (citations removed to ensure anonymity).

Informed by best practices in measurement development,^23^ this multipronged study included two steps: (1) item formulation, and (2) validation in a first sample of primary care clinicians to refine the list of items.

### Item Formulation

The IcanSDM scale aims to assess clinicians’ perceptions of their ability to adopt SDM. An expert panel (AMCG, EF-B, DC) developed a preliminary list of 11 items, in French, based on a review of the literature about the barriers perceived by clinicians to adopt SDM.^19,20,24–31^ Facilitators were not included as they almost all have a barrier counterpart. The panel selected the barriers most often reported and wrote them as affirmative statements (Table 1). Respondents rated the degree to which they agreed with each statement on a visual analogue scale ranging from 0 (strongly disagree) to 10 (strongly agree). We planned to use a total score on the IcanSDM scale from the mean of its items, ranging from 0 to 10 with higher scores indicating higher barrier perception and thus potentially lower ability to adopt SDM.

**Table 1.** IcanSDM items. The items were formulated in French. The English version is a translation that was not culturally adapted.

| **Retained items** |  |
| --- | --- |
| #1 | Shared decision-making results in longer clinical encounters. |
| #2 | Patients often prefer that the clinician make the decision. |
| #4 | Shared decision-making does not apply to all patients, nor does it apply to all clinical situations. |
| #6 | Communicating scientific data to patients is too complex. |
| #7 | Shared decision-making takes up too many resources. |
| #8 | Shared decision-making is inconsistent with clinical practice guidelines. |
| #9 | Shared decision-making is just a passing trend. |
| #11 | *Initial:* The shared decision-making process highlights the uncertainty associated with interventions. This could affect treatment adherence.<br>*Final:* During shared decision-making, the patient becomes aware of the uncertainty associated with interventions and might become confused. |
| **Discarded items** |  |
| #3 | Often, patients have already made their decision. |
| #5 | My team and I already use shared decision-making. |
| #10 | With shared decision-making, I find that many of the interventions I recommend are less effective than I thought. I prefer to continue with my usual practice. |

### Validation

#### Study Participants

In a first validation effort, we recruited a sample of clinicians from various professions (e.g., physicians, nurses, social worker) who worked in family medicine clinics in rural regions of the Province of Québec (Canada). We selected clinics located within a 90-minute drive of Québec City that were not already involved in one of our other studies and thus had never been exposed to the new scale. We initially asked the clinical directors permission to invite the clinicians who practiced in their clinics. If they agreed, we presented the project during one of their scheduled team meetings. After the presentation, the clinicians in attendance were invited to participate in the study, and those who were interested completed the informed consent document and the study entry questionnaire that included questions on their demographic and professional characteristics (e.g. age, profession, year of licensure).

#### Data Collection

Respondents answered electronic surveys before (t0) and after (t1) completion of the web-based training program on SDM. As this was done during a think-aloud session to assess e-TUDE, as described earlier, we also recorded their comments as they completed the questionnaires, but did not prompt them to get their impressions of the items. We transcribed these comments verbatim.

The surveys included the IcanSDM and the Belief about Capabilities subscale of the CPD-Reaction.

The CPD-Reaction questionnaire is meant to measure the determinants influencing adoption of a behaviour, namely intention, social influence, beliefs about capabilities, moral norm, and attitude/beliefs about consequence.^32–34^ The present study reports exclusively the results of the Belief about Capabilities subscale of the CPD-Reaction, which reflect clinicians’ general beliefs about their ability to adopt SDM, and comprises three items, each scored on a 7-point Likert-type scale.

Similarly to the CPD-reaction, IcanSDM is also meant to measures beliefs about capabilities. However, it is more precise than the CPD-reaction as it allows measuring a set of salient beliefs underlying this determinant, as they have been extracted from the literature.

#### Analyses

##### Content validity and item analyses

We analyzed respondents’ comments about each item as they completed the survey, looking for any mention of incomprehension and evaluating acceptance.

For each item, we also visually inspected the distribution of respondents’ responses before and after training to explore each item’s instructional sensitivity.

##### Internal consistency

We evaluated the scale’s internal consistency using Cronbach’s alpha coefficient for measurements made before and after exposure to e-TUDE.

We also checked item-wise consistency using partial correlation coefficients at both t0 and t1.

##### Sensitivity to change

We hypothesized that training primary care professionals in SDM using e-TUDE would increase their perceived ability to adopt SDM, which should result in a lower score on the IcanSDM scale (i.e., fewer perceived barriers). To verify this hypothesis, we compared the means of participants’ total scores before and after e-TUDE using the paired Student’s t test. We also visually compared the frequency distribution of answers to the pooled items before and after training.

##### Convergent validity

To evaluate the convergent validity, we calculated the Pearson’s correlation coefficient (r) between the total score on the IcanSDM scale and the total score on the Belief about Capabilities subscale of the CPD-Reaction. We expected a negative correlation between the two scales.

We conducted all statistical analyses with the SAS package version 9.4 (SAS Institute Inc., Cary, NC, USA). We set the statistical significance of all analyses at 0.05.

### Ethics approval

We obtained ethical approval for the overall project including all phases from the (name of the ethic review board removed, to ensure anonymity).

## Results

### Participant Characteristics

We recruited 16 primary clinicians. Of these, 75% were women and 75% were physicians. They had a mean age of 38 and an average of 10 years of practice (Table 2).

**Table 2.** Characteristics of the participants in the Validation and Confirmation Steps of the study.

|  | **Frequency (%)**<br>**(Total N=16)** |
| --- | --- |
| **Gender** |  |
| Female | 12 (75%) |
| **Age** |  |
| □ 30 | 4 (25%) |
| 30-39 | 6 (38%) |
| 40-49 | 4 (25%) |
| 50-59 | 2 (12.5%) |
| 60-69 | 0 (0%) |
| **Profession** |  |
| Physician | 12 (75%) |
| Nurse | 2 (12.5%) |
| Social worker | 2 (12.5%) |
| Occupational therapist | 0 (0%) |
| Pharmacist | 0 (0%) |
| Physiotherapist | 0 (0%) |
| Nutritionist | 0 (0%) |
| Nursing assistant | 0 (0%) |
| **Years of practice** |  |
| □ 10 | 6 (37.5%) |
| 10-19 | 6 (37.5%) |
| 20-29 | 4 (25%) |
| 30-39 | 0 (0%) |
| Do not recall | 0 (0%) |
| **Area of practice** |  |
| Capitale-Nationale | 0 (0%) |
| Abitibi-Témiscamingue | 1 (6%) |
| Gaspésie—Îles-de-la-Madeleine | 7 (44%) |
| Lanaudière | 7 (44%) |
| Centre-du-Québec | 1 (6%) |

### Content validity and item Analyses

Analyses of participants’ comments revealed that 7 of the 16 respondents (43%) did not understand item #10 (Supplement material 1). One person did not understand the item #11.

To analyze each item’s instructional sensitivity, we did a visual inspection of the histograms of answers before and after clinicians’ exposure to training (Supplement material 2). We first noticed that the frequency distribution of clinicians’ answers to items #1, 2, 3, 4, 5, 8, 9, 10, and 11 shifted towards perceptions of increased ability to adopt SDM (towards 0) after training. In contrast, clinicians’ answers to items #6 and 7 did not consistently demonstrate a positive impact of training, with a small proportion of respondents reporting a decreased ability to adopt SDM (ratings shifting towards 10) after training.

### Internal Consistency

Internal consistency for the initial 11-item scale was low at t0, with a Cronbach’s alpha of 0.19, and increased at t1 with a Cronbach’s alpha of 0.57. Item-wise consistency, using partial correlation coefficients, revealed some problems with negative partial correlations for items #3 and 5 at both t0 and t1 (Table 3). Items #10 and 11 also gave negative partial correlations, but only at t0.

**Table 3:** Item-wise and overall internal consistency as evaluated using the complete list of 11 items.

| Item | Standardized Variables |  |  |  |
| --- | --- | --- | --- | --- |
|  | t0 |  | t1 |  |
|  | Partial correlation | Partial Cronbach's alpha | Partial correlation | Partial Cronbach's alpha |
| #1 | 0.09 | 0.16 | 0.45 | 0.50 |
| #2 | 0.08 | 0.17 | 0.22 | 0.55 |
| #3 | -0.32 | 0.36 | -0.32 | 0.67 |
| #4 | 0.15 | 0.13 | 0.25 | 0.55 |
| #5 | -0.43 | 0.41 | -0.19 | 0.64 |
| #6 | 0.24 | 0.08 | 0.14 | 0.58 |
| #7 | 0.68 | -.21 | 0.34 | 0.53 |
| #8 | 0.42 | -.03 | 0.53 | 0.48 |
| #9 | 0.48 | -.07 | 0.58 | 0.46 |
| #10 | -0.23 | 0.32 | 0.39 | 0.51 |
| #11 | -0.15 | 0.29 | 0.48 | 0.49 |
| **Overall Cronbach's alpha** | 0.19 |  | 0.57 |  |

These findings, as well as those of the previous qualitative item analysis, suggested removing items #3 (*Often, patients have already made their decision)*, #5 (*My team and I already use shared decision-making*), and #10 (*With shared decision-making, I find that many of the interventions I recommend are less effective than I thought. I prefer to continue with my usual practice*) from IcanSDM.

We then re-analyzed our data after these changes, and obtained a Cronbach’s alpha of 0.63 at t0, and 0.71 at t1 (Table 4). Item-wise consistency also improved, though item #11 still presented a negative partial correlation at t0, which might indicate that the item is better understood after training.

**Table 4.** Item-wise and overall internal consistency as evaluated using the selected list of 8 items.

| Item | Standardized Variables |  |  |  |
| --- | --- | --- | --- | --- |
|  | t0 |  | t1 |  |
|  | Partial correlation | Partial Cronbach's alpha | Partial correlation | Partial Cronbach's alpha |
| #1 | 0.13 | 0.65 | 0.60 | 0.64 |
| #2 | 0.15 | 0.64 | 0.24 | 0.72 |
| #4 | 0.16 | 0.64 | 0.31 | 0.70 |
| #6 | 0.45 | 0.56 | 0.16 | 0.73 |
| #7 | 0.76 | 0.46 | 0.32 | 0.70 |
| #8 | 0.61 | 0.51 | 0.64 | 0.63 |
| #9 | 0.58 | 0.52 | 0.57 | 0.65 |
| #11 | -0.09 | 0.70 | 0.43 | 0.68 |
| **Overall Cronbach's alpha** | 0.63 |  | 0.71 |  |

We thus retained eight items for potential inclusion in IcanSDM. The item 11 was also reworded to improve understanding and to address the negative partial correlation described earlier. It was reworded from “The shared decision-making process highlights the uncertainty associated with interventions. This could affect treatment adherence.” to “During shared decision-making, the patient becomes aware of the uncertainty associated with interventions and might become confused.”

### Sensitivity to Change

Before training, respondents reported a mean level of 2.70 (± SD 0.98) in their perceived barriers when items 3, 5 and 10 were excluded. After training, this level decreased to 2.25 (±SD 1.11), indicating a significant 16% decrease in their perceived barriers to adopt SDM (or increase in their ability to adopt SDM; p<0.0001). This shift between t0 and t1 is also apparent in the histogram depicting score distributions (Figure 1).

**Figure 1.**
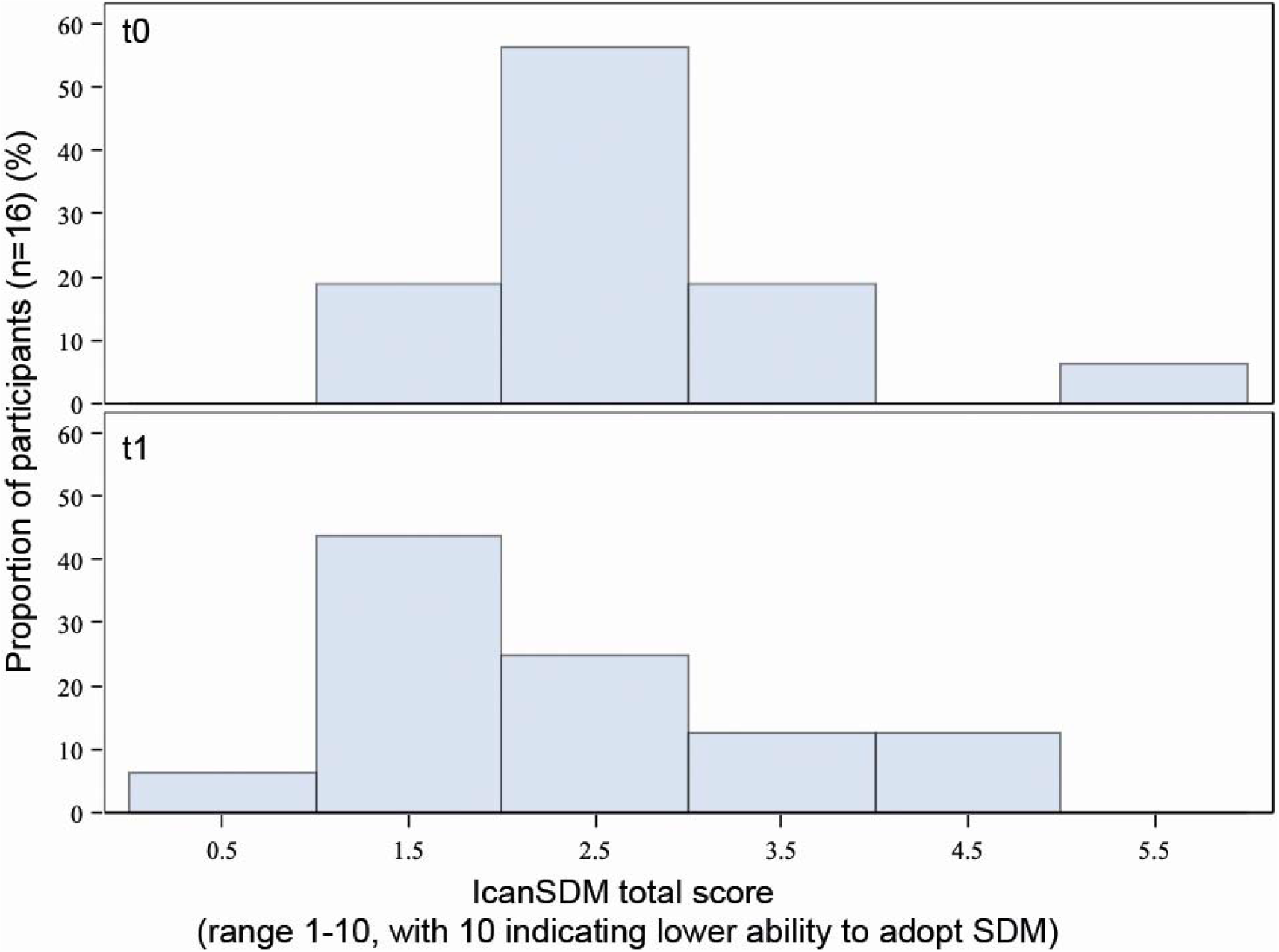
Frequency distribution of answers to the eight items retained, before (t0, top graph) and after (t1, bottom graph) the training program.

### Convergent Validity

We evaluated the correlation between the 8-item IcanSDM and the Belief about Capabilities subscale of the CPD-Reaction instrument. We observed a lack of correlation between the scales at t0 (Pearson’s r = −0.20, p=0.46) and a significant negative correlation between them at t1 (Pearson’s r = −0.59, p=0.02).

## Discussion

We sought to develop and validate a new scale, the IcanSDM scale, to assess clinicians’ perceptions of their ability to adopt SDM. IcanSDM is intended for clinicians of any profession working in any setting, as healthcare is increasingly provided by interprofessional teams.^15^ During the course of this study, the scale was reduced from 11 to 8 items, and one item was reworded. The scale was found to be acceptable to users. Initial validation showed promise of the scale’s ability to indicate impacts of a training program on SDM, as we found acceptable internal consistency, sensitivity to change, and convergent validity. Our results lead us to the following observations.

First, we found that internal consistency of the 8-item IcanSDM version was limited but acceptable (Cronbach’s alpha from 0.63 to 0.71, respectively before and after training), considering this is a new instrument with a limited number of items.^35–37^ Opinions differ about the ideal alpha value. Some experts recommend the alpha should be at least 0.90 for instruments used in clinical settings.^38^ Others suggest an alpha of 0.70 is acceptable for a new instrument^36,37^ and that an alpha higher than 0.90 would indicate repetitive items.^35^

Several scales have been designed to measure SDM processes during the deliberation phase of clinical encounters. A few have also been validated to assess decision antecedents in patients, decision outcomes, or clinician competencies in SDM.^17,18,39^ Some scales are also used alongside an evaluation of SDM processes, to evaluate the impact of SDM on patient health.^40^ However, to the best of our knowledge, no tool, until IcanSDM, has been available to assess clinicians’ perceived ability to adopt SDM. The innovative nature of the scale supports the merits of further testing, as this could help bridge the gap in our ability to understand the determinants of clinicians’ SDM behavior intentions and thus help improve SDM training and implementation efforts. Indeed, SDM behavior is the intended goal of any such effort.

Next steps should comprise testing with a larger sample size that would allow factorial analysis, and a proper analysis of the scale’s sensitivity to change. It would also be interesting to develop a complementary set of items for organizational/contextual barriers to help evaluate implementation efforts more thoroughly.

The strengths of this study lie in the rigorous application of scale development procedures^23^ and the presence of convergent validity testing. The items were based on a review of barriers coming from a large worldwide dataset from diverse populations using various languages. We also evaluated the change in clinicians’ perceptions before and after receiving training on SDM and we led a qualitative exploration of the limits in users’ understanding of the items in order to ensure that IcanSDM helps discriminate effectively between clinicians’ perceived ability or inability to adopt SDM.

Another strength of this study is that the sample included clinicians from various professions (e.g. physician, nurse, social worker). The inclusion of these different healthcare professions support using this scale in various clinical contexts.

Nevertheless, this study also has some methodological limitations. First, there was no control group to measure IcanSDM’s sensitivity to change. Second, the sensitivity to change and convergent validation were based on a relatively small sample size. Further testing with a greater number of users is therefore needed to establish standards to govern the interpretation of results. Finally, the only scale found to test convergent validity—CPD-Reaction—has its own limitations including ceiling effects, although its Belief about Capabilities subscale shows the highest sensitivity to change.^34^

## Conclusion

The 8-item IcanSDM scale assessing clinicians’ clinical behavioral intentions concerning SDM showed promising validity and reliability. Further testing is required to confirm its responsiveness to changes. To our knowledge, there is currently no tool available to assess clinicians’ perceived ability to adopt shared decision making during clinical encounters with patients, despite the increasing worldwide interest in implementing this approach. Indeed, the current shared decision-making research agenda is heavily invested in studying implementation of shared decision making, as the barriers to implementation appear very challenging to overcome.^10^ A validated quantitative measure of clinicians’ perceived barriers is thus needed to help optimize interventions and to assess how our training programs are able (or not) to change clinicians’ perceptions of these barriers. IcanSDM could also be used to identify the clinical settings where clinicians perceive themselves as being least able to adopt shared decision making, as part of a training needs assessment.

## Data Availability

The datasets used and/or analysed during the current study are available from the corresponding author on reasonable request.

## Funding sources

This work was supported by the *Ministère de l’Économie, de l’Innovation et de l’Exportation du Québec*, and by the *Société de Valorisation SOVAR* (grant number 2014-2015-PSVT2-31494). We also received in-kind support from the Office of Education and Continuing Professional Development of Laval University and *Centre de recherche sur les soins et les services de première ligne de l’Université Laval (CERSSPL-UL)*. AMCG is funded by a Research Scholar Junior 2 Career Development Award by the *Fonds de Recherche du Québec—Santé*.

## Acknowledgements

We would like to thank Émilie Fortier-Brochu (EF-B) and Danielle Caron (DC) for their participation as experts to the panel to formulate the items list. We would also like to acknowledge the contributions of Danielle Caron (DC) and Moulikatou Adouni Lawani to data collection and analysis. We also wish to thank Katherine Hastings for the writing assistance.

## Author Contribution

AMCG designed this study and led the expert panel in charge of creating the items list. AMCG and LC collected the data. AMCG, P-HC, JSR, LC, and LB analysed, and interpreted the data. AMCG, LC, and LB participated in the initial drafting of the manuscript. All authors drafted, critically revised and gave final approval of the article. AMCG acts as guarantor.

## Ethics approval

We obtained ethical approval for the overall project including all phases from the Ethics Review Board of the *Ministère de la Santé et des Services Sociaux* (reference CCER15-16-05).

## Supplementary material

**Supplement 1**. Participant comments made while completing the IcanSDM scale.

**Supplement 2**. Frequency distribution of answers to each item before (t0, top graph) and after (t1, bottom graph) the e-TUDE training program in the study group.

## Notes

### Competing Interest Statement

The authors have declared no competing interest.

### Funding Statement

This project was funded by the Ministère de lèÉconomie, de l’Innovation et de l’Exportation du Québec and by the Société de Valorisation (SOVAR) (grant number 2014-2015-PSVT2-31494). We also received in-kind support from the Office of Education and Continuing Professional Development at Laval University and from the VITAM Research Centre on Sustainable Health. AMCG is funded by Research Scholar Junior 2 Career Development Award by the Fonds de Recherche du Québec-Santé. LB is funded by fellowship from the Canadian Institutes of Health Research. FL is funded by a Tier 1 Canadian Research Chair in Shared Decision Making and Knowledge Translation. The funders had no role in study design, data collection and analysis, decision to publish, or preparation of the manuscript.

### Author Declarations

We obtained ethical approval for this project from the Ethics Review Board of the Ministre de la Santé et des Services Sociaux du Québec (reference CCER15-16-05) and the Centre Hospitalier Universitaire de Québec (reference 2016-2521). All participants signed an informed consent prior to participation in the study.

